# Navigating online information access for women survivors of intimate partner violence living in long term shelters

**DOI:** 10.1101/2024.12.02.24318365

**Authors:** Ebony Rempel, Lorie Donelle, Jodi Hall

## Abstract

This study explores the use of online resources by women who experienced intimate partner violence (IPV) and were living in second-stage shelters. Given the ubiquity of online access across all aspects of everyday life—from health care and education to job searching and social support—ensuring equitable digital access is essential for everyone. This study used purposive sampling and thematic analysis of in-depth, in-person interviews with women residing in second-stage shelters across Alberta, Canada, to explore their experiences with online resources for support and information. Thematic analysis identified three main themes: Proactive Preparation, Staying Connected to Support Networks, and Barriers to Online Access - highlighting the critical role of digital resources in empowering participants but also underscoring significant challenges, such as financial constraints, internet reliability, and privacy concerns. Participants emphasized the importance of online resources for maintaining relationships, preparing for meetings with service providers, and accessing information and support. However, they faced significant challenges, including financial constraints, lack of reliable internet access, and privacy concerns. The findings underscore the need for improved digital access, health equity, and tailored digital literacy programs to support IPV survivors effectively. While social media and online platforms provide vital support and information, they also pose risks of digital surveillance and stalking. The study advocates for a collaborative effort from government agencies, service providers, healthcare providers, technology companies, and community organizations to create comprehensive support systems. Addressing these barriers can enhance the accessibility of crucial information and resources, empowering women on their journey towards recovery and independence. Introduction

**Author Summary:** The researchers shed light on the experiences of women who have experienced IPV who seek information and support through online resources while residing in second-stage shelters. Recognizing that digital access has become a staple of modern life, our research investigates how these women navigate online spaces to support their journey towards recovery. Through interviews with women across Alberta, Canada, we identified critical themes: the need for proactive information gathering, maintaining connections with support networks, and the challenges posed by limited online access. Participants spoke to the value of digital resources for maintaining relationships and preparing for important interactions with service providers, while also facing significant barriers like financial constraints, unreliable internet, and privacy risks. Our findings call for collaborative efforts from service providers, policymakers, and technology companies to improve digital accessibility, privacy safeguards, and tailored literacy programs. By addressing these obstacles, we aim to empower women in second-stage shelters, helping them build self-efficacy and resilience through secure and supportive online environments.

## Introduction

Navigating the multifaceted dynamics of IPV constitutes a significant challenge for women globally. From the subtle manifestations of coercive control to the overt expressions of physical abuse, each woman’s trajectory through IPV is inherently unique, characterized by emotional upheaval and often requires them to make life altering decisions [1]. Within this context, access to information, essential services, and support assumes importance.

The information journey embarked upon by women navigating IPV is often fraught with challenges and uncertainties. Initially, many individuals grapple with recognizing and understanding the signs of abuse within their relationships, confronting societal norms and misconceptions that may trivialize or normalize such behavior [2,3]. As they seek clarification and validation of their experiences, women who have experienced violence encounter a labyrinth of resources, ranging from offline (e.g. pamphlets and educational materials) to online resources (e.g., safety apps, discussion forums, and helplines) [4]. The internet has become an indispensable tool for accessing vital information and support services, enabling women to prepare for interactions with healthcare, legal, and social services professionals. Online resources, including safety apps and social media platforms, provide discrete avenues for survivors to seek help, connect with supportive networks, and gain knowledge to navigate their unique circumstances effectively [5]. The increased use of online platforms and access to digital resources play a crucial role in the lives of women survivors of IPV residing in long-term shelters [1,5]. However, the reliability and comprehensiveness of relevant and helpful information may vary significantly, challenging one’s access to information (e.g., readability of information and literacy skills) and complicating the process of readers’ ability to discern accurate and reliable information from false[6]. Moreover, cultural and linguistic barriers may exacerbate the difficulty of accessing pertinent online resources, particularly among marginalized communities [1] adding to the information and resource access challenges among online information seekers and especially among women who have experienced IPV.

### Shelters for those who have Experienced Violence

Shelters for those who have experienced domestic violence, often referred to as domestic violence shelters or women’s shelters, play a crucial role in providing safety, support, and resources for individuals who are experiencing violence in their relationship(s) [7]. Emergency shelters offer short term accommodation to women and their children, providing temporary refuge from their abusers. Second-stage shelters offer longer-term, supportive housing, often for those transitioning from emergency shelters to independent living. The primary goal of emergency domestic violence shelters is to offer immediate protection and support to individuals experiencing abuse, while second-stage shelters assist them in accessing long-term solutions and resources to rebuild their lives [8]. Second-stage shelters, also known as transitional shelters, provide extended, supportive housing for women who have experienced violence. These shelters offer an environment where women can reside for several months to a few years, allowing them time to heal and rebuild their lives. Second-stage shelters often provide access to diverse resources ideally aimed at helping women achieve independence from their abusive situations and long-term stability [9].

Shelters offer a range of information and support services tailored to the unique needs of women who have experienced violence. These services may include temporary accommodation, crisis intervention, counseling, legal advocacy, and assistance with accessing community resources such as healthcare, childcare, and housing assistance [10]. Many shelters also offer support groups and educational workshops to support survivors and help them navigate the complexities of abusive relationships [10]. Accessing emergency and/or second-stage shelters can be a crucial step towards safety and healing for those who have experienced violence. Shelters typically have 24-hour hotlines staffed by trained professionals who can provide immediate support, information, and guidance to individuals in crisis. Additionally, shelters often collaborate with local law enforcement, healthcare providers, and social service agencies to ensure survivors receive comprehensive support and assistance.

Overall, domestic violence shelters play a vital role in helping women and their children escape abusive situations and rebuild their lives [10]. By providing temporary accommodation, support services, and resources, shelters offer women temporary refuge from violent partners, and support women to pursue a life path free from the cycle of violent and abusive partners.

### Online Information Access

Women aged 15 to 24 years are at the highest risk for IPV [11–14], and are also some of the most prevalent users of the internet, online technologies, and digital devices [15,16]. Historically, women who experienced IPV reported a preference for face-to-face and over the phone communication and information [17]. However, women of IPV are now accessing and benefiting from the availability, ease, and convenience of online resources [18]. The societal acceptance of digital tools and the limitations of IPV support systems are influential factors facilitating online information seeking among women who have experienced IPV [19]. However, the use of digital technology to surreptitiously track online behaviours and activities of others creates an element of danger for women who have or have left abusive partners [20].

There is a growing need to understand the use of technology to support access to information, services, and support among women who have experienced IPV to match the increased need for specialized and supportive IPV related resources such as shelters, justice system, online supports, and counselling services [21]. A recent scoping review of online interventions for women experiencing IPV revealed that interventions focused on leaving the abusive relationship with limited attention to post-relationship experiences and broader social-contextual factors [22]. This research underscores the need for comprehensive support to extend beyond the point of leaving the abusive partner or situation and indicates gaps in long-term needs of survivors accessing online information and services [23]. Similarly, a US nationwide analysis of online or e-government support for people who have experienced IPV found that general crisis related resources (e.g., hotline or referral) were more prevalent than content related to specific needs (e.g., child custody); for women who have experienced IPV, there was more information on emergency needs (e.g, creating a safety plan, social support) compared to long-term needs (e.g., housing, adequate income, childcare services) [24–29].

Smartphone applications have become indispensable tools of daily living, including addressing issues as sensitive as IPV [5]. Designed with user safety and support in mind, many online applications offer discreet avenues for individuals experiencing IPV to seek assistance, access resources, and document incidents securely [5,30]. For example, the myPlan app (https://myplanapp.ca/) is an interactive app that helps individuals assess relationship safety and create a personalized plan to stay safe, providing resources and guidance for those experiencing or concerned about relationship abuse. Those who download the app can input specific information (i.e., urban or rural area, number of children) and the app will provide an assessment creating a safety plan that gives women time to think about their choices. Assuming adequate literacy and digital literacy skills, the availability of online apps and resources such as emergency hotlines, chat support, safety planning tools, and information on legal rights, facilitate access to information among IPV survivors and the opportunity to connect with the help they need, using their smartphones [5]. In an era where technology increasingly intersects with social challenges, these applications represent a crucial step forward in leveraging innovation to support those affected by IPV [25,31].

Social media sites are online web-based networks where individuals and corporations can create a profile, and connect with other users via text, video, and pictures [15]. Researchers evaluated the mobile app, *SafetiPin*, that crowd sources data and collects information on women’s safety in India as an effective way to provide data to users and stakeholders as a means of increasing awareness and activism in the VAW sector [32]. SafetiPin’s crowdsourced approach has empowered users and stakeholders by providing real-time safety insights, fostering increased awareness, and supporting activism within the VAW sector, thereby illustrating the potential of mobile technology to create safer public spaces for women. However, there is limited research on how newer forms social media networks (e.g., Facebook and Twitter) and smartphone applications, are used in terms of accessing information, especially among women experiencing IPV [19,33,34]. The rapid pace of development in digital technologies makes it nearly impossible to continuously track and evaluate emerging online tools, as new platforms and applications are introduced at an unprecedented rate, often outpacing efforts to assess their effectiveness and safety for users.

Here’s a possible introductory sentence for your article:

In a study that explored the role of online resources in supporting women survivors of IPV residing in second-stage shelters, researchers reported differences in service provider technology readiness [35]. The authors concluded that more research is needed to enhance provider technology readiness and to understand which types of technology (e.g., mobile apps, online resources, and other digital tools) are helpful for those who have experienced trauma related to IPV. Within this ‘information age’, service providers’ digital health literacy, knowledge and skill is essential in providing contemporary services to women at critical life junctures.

While insight into online resources (e.g, safety apps) for women experiencing IPV is growing, relatively little is known about online information resources and information needs among women living in second-stage shelters. There has been limited research attention given to understanding the processes of accessing information, services, and supports and whether and how women living in second-stage shelters are accessing online spaces [2,10,36,37]. Women’s decision to leave an abusive situation and seek shelter services often necessitates interaction with the multiple health (e.g., medical services), social (e.g., income, housing, etc.), and legal systems that requires considerable time and effort to coordinate [38–41]. As technology continues to evolve, it is imperative to address these research gaps to ensure that all women, regardless of age or circumstance, have equitable access to the online information and support they need to navigate and overcome the challenges of IPV. With digital devices now ubiquitous, many organizations—particularly nonprofits—are leveraging online-only resources to increase accessibility and achieve fiscal efficiency, enabling them to serve a broader audience with limited resources [42]This shift highlights the necessity for comprehensive digital strategies that can sustain and expand service delivery in resource-constrained environments. The research question that guided the overall research was: How do the information journeys of women living in second-stage shelters in Alberta reflect their evolving needs for support, services, and information? This manuscript is part of a larger study investigating the information, services, and support needs of women experiencing IPV who are accessing second-stage shelters. The broader research aims to understand the comprehensive journey of these women as they navigate various systems and services. Specifically, this research is a secondary analysis [43] of data from the larger study focusing on the women’s information journey within the online or digital context, exploring how digital tools and resources support their search for knowledge and safety in the face of IPV. An investigation of the information, supports, and service needs of women who access VAW second-stage shelters will contribute to the limited evidence available to service organizations, health care providers, and policy makers in effectively responding to the needs of women who have experienced IPV and its consequences.

## Methods

A detailed reporting of the research methods are available elsewhere[44]. Women residing in the province of Alberta, Canada, who were living or had previously lived in any of the 14 provincially funded second-stage shelters in 2019 were recruited for the study. Recruitment efforts involved distributing flyers to all shelters across the province and directly engaging with executive directors for collaboration. Interested participants contacted the researcher directly, received study information, and confirmed participation; attention was given to recruiting participants from diverse geographical areas within the province [45].

Purposive sampling was used to identify women using the following inclusion criteria: (1) 18 years of age or older, (2) self-identified as women, (3) currently lived in a second-stage shelter in Alberta or had lived in a second stage shelter within the past 12 months, and (4) could speak, read, and write English. The number of participants was determined based on the richness and depth of data collected. The initial coding process involved dividing participants’ stories into smaller segments, each highlighting various parts of their experiences in the shelter, such as difficulties in finding information and support. As recurring patterns appeared, more participants were added to examine elements of shelter life until all category characteristics were fully explored. Data collection concluded once no additional codes emerged. Each participant received a $25 honorarium for their time and any associated costs.

Data collection took place through in-person interviews in locations that were mutually agreeable to the participant and the researcher. Interviews lasted an average of 64 minutes. During analysis, each interview was compared to previous ones to refine categories based on similarities and differences in participants’ experiences. The lead author conducted line-by-line coding, focusing on the question, “what does this data suggest?” Initial coding identified common experiences across participants, highlighting a dominant social process. Axial coding linked the main category of ‘navigating the shelter system’ to subcategories like ‘information-seeking behaviors’ and ‘barriers to accessing support.’ Through selective coding, the emerging theory of ‘navigating the maze of shelter systems’ was developed, integrating subcategories related to accessing information and support. Ethical approval was obtained from Western University’s Non-Medical Research Ethics Board.

### Findings

Twenty women residing in Alberta, Canada participated in the study. The average age was 37.6 years (ranging from 25-61 years). The average annual income was $14,954 (ranging from $0-$50,000). The self-reported source of income for participants was mostly government income support assistance and child tax benefit, two participants received income from employment. Education attainment of participants ranged from some high school to university degree. All the participants had children, with two as the average number of children. Seven participants lived in rural areas and 13 lived in urban areas of the province. Eight women identified as new immigrants to Canada, five women identified as First Nation, three women identified at Métis, and four women identified as Caucasian Canadians. Prior to their residence at the second-stage shelter, 15 women lived in an emergency shelter, four lived with their ex-partner and one woman lived with her family. Eighteen participants were currently living at a second-stage shelter, one was in the process of moving out and one was living on her own in the community.

Participants’ use of online resources was summarized into three themes with associated subthemes: 1) Proactive Preparation with subthemes of: Engaging with Social Media for Information and Support and Information Overload; 2) Staying Connected with the subtheme of Social Media Support; and 3) Online Accessibility with associated subthemes of Hardware Access and Connectivity and Privacy Concerns.

### Proactive Preparation

Participants described accessing online information to enhance their knowledge and build their confidence in preparation for appointments and meetings with a wide variety (e.g., health care, legal, employment, education) of professionals. Women sought online information to prepare for meetings with service providers, to familiarize themselves with information of relevance to their current situation and to the system representative (e.g., lawyer) stating “you don’t know what you don’t know” (P13). Participants used search engines like Google to identify relevant resources and information. This proactive approach to meeting with health, social, and other services allowed them to enter discussions with professionals armed with a basic understanding of information relevant to their current situation, enabling more informed interactions and potentially facilitating more effective support and assistance. One participant highlighted the usefulness of search engines like Google in discovering resources and information they were previously unaware of:

But there is lots of help. Just go to Google. You can find it. This is things I didn’t know. (P6)

We live in an online world and there are times when going online is the only option and women indicated how important the ability to go online was to them.

> Even filling out forms and stuff like that and yeah, I wouldn’t be able to live without [the internet]. (P12)

Participants extended their online exploration beyond immediate meeting preparation, using the internet as a tool to access a diverse array of information. As part of their online information-seeking journey, participants also discussed utilizing versatile platforms that offered features beyond traditional web browsing. For instance, one participant highlighted an online Smartphone application she turned to for information:

> It’s an app, but also a website so they send you newsletters and it’s a baby tracker too so when you were pregnant it tracks you. (P5)

English was not the first language of several participants in this study and one participant discussed a way they used the internet to support them in seeking and accessing (translation services) online information:

> I use Google translation [as a] start, [after] my mom left, [it is Google Translate] that I start from. (P6)

### Engaging Social Media for Information and Support

Social media platforms played a crucial role in participants’ search for knowledge and support. They utilized platforms such as Facebook, Reddit, and LinkedIn to access diverse resources, seek advice, and connect with online communities. Despite the burden of information gathering, participants valued the supportive networks and resources available online, highlighting the multifaceted nature of their online interactions.

> Oh, yeah. I, yeah. I would be lost without it. 100%. And even like on Facebook messenger, I have like a group of women that I now say, does anybody have a cheese grater that they’re not using anymore? Boom. One shows up kind of a thing. So, oh, yeah, that’s been a lifesaver for sure (P12)
>
> I also use Facebook groups and then some law groups for Facebook and I put my story out there. Reddit, I’ve been on that. LinkedIn, Twitter. I’ve gone on all of them and put my entire story out there and asked for legal advice. (P15)

### Information Overload

Social media constituted a significant component of online activity among participants. However, women also shared the burden of having to seek and gather information in preparation for meetings with others and to inform the many everyday decisions created by their unique circumstances. The quote that follows expresses participants appreciation but also feelings of burden related to information seeking and gathering.

> I’d have to think back because I think sometimes I’m overwhelmed or I think I’d rather just ask this person first instead of looking it up because it does, it takes time and mental energy to look things up and if someone else can answer to me a lot more smoothly, sometimes I do go that, but yeah, sometimes I’m curious and if I don’t find the answer, yeah, I’m gonna look into it and then ask them. (P16)

In the context of seeking information and support online, the process is often multifaceted and dynamic and as one participant described, this may involve posing questions, seeking advice, sharing personal experiences, and offering support to others within online forums or social media groups dedicated to relevant topics such as qualifications for shelter services as described here:

> there was this woman who was in a domestic violence marriage and she escaped … I remember reading her story and she was talking about going to a shelter and at the time the shelter felt like – well, I wouldn’t qualify and just reading her long post, it was like wow, the shelter is for anybody. (P16)

### Staying Connected to Support Networks

Often women were required to leave their city or neighbourhood to escape an abusive relationship. Keeping in touch with support networks and finding new support networks was important for many participants.

> It’s kind of a little bit lonely but the few relationships I had were maintained by Facebook and I really appreciated them more so because I can’t have somebody in my realistic life every day. (P18)

### Maintaining Support Networks through Social Media

Participants who had to leave their communities to escape abusive relationships emphasized the importance of staying connected with supportive family and friends through social media platforms like Facebook. Despite physical distance, participants maintained their relationships through online platforms, highlighting the role of social media in mitigating feelings of loneliness and isolation. They talked about accessing social media for information, and to connect with family and friends.

> Yeah, my mom and my brothers and my sister. It’s [social media] just the best way to contact each other is through Facebook because sometimes my mom doesn’t have minutes [limited phone plan] or especially where they live. They live in the middle of nowhere so they don’t have cell service a lot, but they’ll have internet, so yeah. (P5)

### Online accessibility

Participants discussed various challenges that hindered their access to online information, services, and supports.

> I can look up [what I need online] myself on my cell phone… But sometimes it’s hard because not everything I need is available without paying, and I can’t always afford it. Plus, with the stalking stuff, I don’t always feel safe using my phone freely. (P8)

Factors such as cost, device access and/or ownership, and the risk of online stalking curtailed their ability to utilize online resources effectively.

### Hardware access and connectivity challenges

Challenges related to internet and cellular service costs, unreliable access to devices, and privacy concerns within shelter settings further compounded participants’ difficulties in staying connected and accessing online support. Some shelters did provide access to WiFi and to devices (e.g. computer). However, many participants also reported that they were accountable for their own internet and cellular service costs; these were not provided services with second stage shelter accommodations.

> [Access to the internet is] really important, really important. Here in the shelter, we don’t have internet service and I’ve been finding that, it’s like my hands are tied behind my back. I can’t afford internet. I just can’t. Between you know, I just can’t and it’s outrageously expensive. So… what I do is in the evening around six thirty, I get in my car. I drive around, I sit in the [Tim Hortons] parking lot and then I can get internet service. (P20).

And another participant conveyed a similar situation stating.

> We don’t have [internet services] – no, each person is responsible for own WIFI. (P14)

The cost challenges of online services are further complicated by unpredictable access to devices like computer equipment. Even though computers stations were included in some of the second-stage shelters, participants reported instances when the equipment was not functional, or the lack of privacy (public space) prohibited use:

> Suddenly, they say [the computer] broken, broken, broken and they take it out, the four computer[s] and they make one computer in small room. (P19)

### Privacy Concerns

Participants expressed concerns about maintaining privacy while accessing digital resources and support services. They noted that service providers frequently required extensive personal information to be documented online, ranging from financial details and housing needs to personal histories with family, healthcare, and legal issues. This level of transparency often left them feeling vulnerable, as some felt pressured to disclose more than they were comfortable with to gain access to essential support. Women may want to be private, yet service providers often require women to show them everything, you get nothing until they know everything about you. This constant sharing of personal details felt invasive to many, leading to a sense of overexposure and vulnerability. One participant noted:

> You know, and it was kind of this wake up call like yeah, you’re probably a lot more visible than you realize and it’s made me really nervous. (P16)

For some, this heightened visibility, especially within digital systems, intensified anxieties around privacy, reinforcing the delicate balance between accessing help and maintaining control over their own stories and personal boundaries. The information required made many participants uncomfortable, as they had to reveal intimate details of their lives. One participant described their strategy of concealing as much as possible online to protect their privacy. They felt compelled to ‘disappear’ online, avoiding certain platforms and minimizing their digital footprint to remain hidden.

> And so, it was just a matter of hiding as well as possible and disappearing and so, once a few months went by then I started thinking, oh well maybe I’ll – no. (P9)

This quote illustrates the constant vigilance and hesitation around maintaining privacy in a world where accessing help often meant sacrificing parts of it. The constant fear of being tracked or monitored by an abusive partner further complicated their use of digital tools, as they had to balance seeking information and support with the risk of compromising their safety. One participant described the realization of how visible she was online, which heightened her anxiety and made her more cautious about her digital footprint. This need to be (in)visible created a paradox where the need for privacy conflicted with the necessity of seeking support, leading to feelings of nervousness and apprehension.

> Because he was stalking me… I ended up having to call the police … they looked at my social media… Facebook. my settings were pretty visible… it made me really nervous. (P8)

Especially women who have left abusive partners, have heightened online privacy concerns potentially impacting their ability to utilize online resources. This creates an additional layer of stress and difficulty in their journey toward recovery and empowerment. The need for privacy and discretion in digital interactions underscores the importance of developing secure and confidential online environments for women experiencing IPV.

## Discussion

This manuscript is part of a larger study on access and use of information, services, and support needs of women living in second-stage shelters who had experienced IPV. Attention to women’s online information seeking and the critical role of digital access and technology readiness, warranted a focused exploration [43]. The thematic findings highlighted participants’ use of online resources and were categorized into three main themes: Proactive Preparation, Staying Connected to Support Networks, and Online Accessibility. The data from the research also underscore issues of health equity related to access to devices, payment for internet services, and health literacy/digital health literacy skills. These findings illustrate the complex interplay between online resource utilization and the challenges faced by women experiencing IPV, emphasizing the need for improved digital access, health equity, and privacy protections in support services. Employing a grounded theory, women participants who lived in second-stage shelters described the importance of online resources for developing and maintaining relationships, to support their knowledge and in preparation to meet with service providers, but also shared their unique challenges specific to online access to information, services, and support.

### Digital Empowerment vs. Cognitive Burden

Unquestionably, widespread services and resources are moving to online formats and given the proper support and information literacy, the internet can be an empowering space for those who have experienced IPV to access information. The lack of affordable and reliable internet access and digital devices disproportionately affects marginalized groups, exacerbating existing inequities [46]. The study “Internet Access and Use in Domestic Violence Shelters: Policy, Capacity, and Management Barriers” by Westbrook (2013) examined the challenges faced by domestic violence shelter administrators in integrating Internet resources into their services. The research was conducted in two stages: a mixed-methods questionnaire to shelter managers across Texas, and a content analysis of 65 shelter websites [47]. Although shelter administrators recognized the potential of the Internet to increase staff efficiency (e.g., streamlining case management and resource referrals) and enhance clients’ sense of self-efficacy (e.g., accessing job training materials and online support networks) but struggle with limited administrative support tools and cyber-safety resources. The majority of shelters lacked formal internet policies and sufficient training for staff and clients. Only a small percentage of the websites included essential cyber-safety information, despite significant concerns about online stalking. The researchers emphasizes the need for a user-centered approach to Internet integration in shelters, considering clients’ expectations and the realities of available resources [47].

A diversity of communication strategies such as face-to-face and the use of online formats are used by women experiencing IPV. The use of mobile digital devices and online communication has become widespread within the public and also among women experiencing IPV [5]. Tarzia and colleagues (2018) interviewed women who had experienced IPV and found that they valued the support received online and that online interventions (e.g., websites and mobile apps) raise awareness about IPV through interactive tools and help reduce feelings of isolation by connecting women to others with similar experiences [48]

Researchers found that women who had experienced IPV valued several aspects of face-to-face support—such as awareness-raising, personalized support, and tailored recommendations—which can also be effectively delivered through online platforms [48]. In a study of 259 participants, including 101 women with IPV experiences, researchers investigated the factors that influence women’s acceptance and use of online domestic violence resources [42]. The findings from this study emphasized the need to understand everyday information-seeking behavior as necessary for designing effective support sites; considering user needs, emotional states, cognitive factors, and available resources to ensure the sites are comprehensive and user-friendly. The findings from this research highlighted the importance of creating accessible, up-to-date, and relevant online resources to support IPV survivors effectively [42]

### Social Media as a Double-Edged Sword

We theorize that social media can play a paradoxical role in the recovery journey of IPV survivors. Social media platforms provide significant benefits for women experiencing IPV, including access to information, connection with others who have similar experiences, and support from virtual communities. Social media platforms can facilitate rapid dissemination of information, allowing users to access a wide range of resources and perspectives tailored to their needs [49]. However, the same features that make social media useful can also pose risks especially to women and more so among women who have experienced IPV [50]. Women often face the burden of managing their online presence to avoid digital surveillance and stalking, adding stress to their already challenging situations. Despite these challenges, social media remains a crucial tool, enabling women to navigate their everyday lives with greater agency and resilience. Programs like Women’s Shelters Canada’s Technology Safety Canada project provide essential resources and training to mitigate online stalking, offering practical safety tips and virtual training for survivors and anti-violence workers. Despite the educational benefits of programs like Women’s Shelters Canada’s Technology Safety Canada project, which aim to mitigate online stalking, a disproportionate burden remains on IPV survivors to protect their own digital security; while these initiatives raise awareness and offer immediate support, they often shift responsibility onto women to manage their safety by limiting their online presence and relying on privacy tools. The onus of protection is often placed on those already vulnerable, rather than addressing the root causes of digital surveillance and ensuring that safer, more secure online environments are created and maintained by service providers and technology platforms.

For women managing IPV, social media can be both helpful and harmful; it serves as a vital space for connecting with supportive communities and accessing tailored information, yet it also exposes women of IPV to risks of digital surveillance by perpetrators. Online stalking and technology-based abuse present unique risks for IPV survivors, complicating their recovery journeys. As researchers describe, technology-based abuse enables abusers to maintain control over survivors through constant digital surveillance, harassment, and coercive tactics, creating a sense of omnipresence that can make leaving an abusive relationship feel nearly impossible [50]. Other research also highlights how perpetrators exploit tools like GPS tracking, spyware, and social media, leveraging these technologies to monitor survivors’ movements, access their personal information, and disrupt any sense of safety [51]. These forms of digital intrusion often blur boundaries between online and offline abuse, as the perpetual risk of surveillance instills continuous fear and anxiety, affecting survivors’ ability to engage safely with online resources [50,51]. Such risks reinforce the need for comprehensive support systems and emphasize that, while social media and online resources offer valuable support and community, they can also expose survivors to heightened dangers of digital stalking and abuse. Addressing these barriers requires collaborative efforts from government agencies, service providers, healthcare providers, technology companies, community organizations, and academic institutions to create comprehensive support systems that empower IPV survivors.

### Socioeconomic Barriers to Digital Inclusion

A recent study by Micklitz and colleagues (2023) explored the acceptance and usability of digital interventions for IPV survivors and found that digitally adapted interventions could address the needs of those who have experienced IPV by reducing social isolation, facilitating self-reflection, and offering coping strategies [19]. The study involved six individuals with lived IPV experiences and six service providers, using the think-aloud method and semi-structured interviews. It found that digital self-help tools can reduce social isolation, provide space for self-reflection, and offer coping strategies. However, challenges such as ongoing violence, varying psychological capacities, and different stages of awareness among survivors complicate the development of effective digital interventions. The study emphasizes the need for a multi-modular, trauma-informed digital intervention with appropriate security measures to support IPV survivors comprehensively [19]. Women in the current study reported several barriers to online use that included the overwhelming amount of information available online, the onus of preparing for meetings, issues with access to hardware and a Wi-Fi connection, and a concern for the online privacy. Financial constraints and lack of access to reliable hardware and internet connectivity create substantial barriers to the effective use of digital tools. The limited financial resources typical among women experiencing IPV create substantial obstacles to accessing reliable internet or cellular services, which are especially costly in Canada, where internet and cellular rates rank among the highest globally [52]. These costs place a heavy burden on survivors who, without stable or affordable connectivity, may lack access to essential mobile support and real-time assistance when they need it most. This barrier becomes even more concerning given the heightened risks of online stalking and abuse; survivors often navigate restrictive digital practices to protect themselves, including limiting their online presence or managing complex privacy settings, which can feel isolating and unsafe without proper guidance.

In addition to the personal financial constraints on survivors, shelter organizations also face resource limitations that affect the support they can provide. Many shelters operate on limited budgets, which can impact their capacity to offer devices, consistent internet access, or cyber-safety tools necessary for survivors to safely access digital resources [53]. Furthermore, without formal policies, administrative resources, or training on internet use, shelter staff may lack the digital literacy and technical knowledge to assist survivors effectively. This gap means that even when internet access is available, support for navigating online resources and ensuring digital safety can fall short, potentially leaving survivors without sufficient digital support to navigate their recovery journey. As Westbrook (2013) emphasizes, shelters express a critical need for comprehensive administrative tools, including internet use policies and training materials, as well as targeted resources to address cyber-safety concerns like online stalking and abuse. Addressing these gaps could significantly improve the quality of support shelters offer, creating a safer and more empowering digital environment for survivors.

Cyber-safety is a critical concern across these groups, with significant risks of online stalking and abuse [54]. Despite these challenges, internet access can enhance self-efficacy and provide vital social support, helping individuals make informed decisions and maintain social connections [55]. A user-centered approach to integrating digital tools, focusing on clients’ needs and expectations aligns with broader literature advocating for responsive and inclusive digital infrastructures for marginalized populations [56].

To address these barriers, tailored digital literacy programs, accessible online resources, community support networks, and awareness campaigns promoting privacy and security are essential [57]. Addressing barriers to digital access for women who have experienced IPV requires a collaborative effort from government agencies, service providers, healthcare providers, technology companies, community organizations, and academic institutions. By tackling these obstacles, we can enhance the accessibility of crucial information and resources for women who have experienced IPV.

Women’s shelters like other organizations are increasingly creating online websites [58]. Information about the organizations’ mandate, administrative resources, services, and operating hours, are publicly available to those who have the knowledge (literacy), skills (digital), and resources (devices, internet provider) to access the information. While some shelters provide access to online resources and information, there is limited information available regarding what this access looks like (e.g., devises to access and use, WIFI, training, and support to access online information). Further research is needed to understand what is available for those accessing shelters and what the needs are for online information access. There is limited research available on the differences that marginalized groups (e.g., older adults, newcomers) may need in terms of online information support.

This research has significant implications for practice and policy. There is a need to enhance technology readiness and digital access among IPV survivors, service providers, policymakers. It is important that IPV survivors, service providers and policy makers work collaboratively with technology developers to mitigate barriers to use and create opportunities for financial support for internet access, digital literacy training, and development of secure platforms that protect users’ privacy and safety.

### Limitations

Only English-speaking participants were invited to participate in the study. Women who had English as their second language participated and though translation services were offered via an over-the-phone translation option, no participants opted for this option. Participants were recruited from across a single province within Canada and women from other provinces or territories within Canada may have different levels of access to second-stage shelter services. These limitations provide a motivation for future research.

## Conclusion

Given the ubiquity of internet and digital device use, online services, information, and support are a public necessity and uniquely relevant women who have experienced IPV. Women go online to access information, service, and support and for formal and informal support. There is recognition that some information is only available online and that those who have experienced IPV may choose to access information online and therefore shelters should continue to make the move towards offering services and providing access to online options for information, services, and support.

## Data Availability

The data supporting the findings of this study were collected through in-depth, in-person interviews with women residing in second-stage shelters across Alberta, Canada. Due to the sensitive nature of the data, including personal and potentially identifying information about participants who are survivors of intimate partner violence, the data cannot be made publicly available to protect participants' confidentiality and safety. Requests for access to de-identified data for research purposes may be directed to the corresponding author, Ebony Rempel, at. Access will be considered on a case-by-case basis, in accordance with ethical guidelines and subject to approval from Western University's Non-Medical Research Ethics Board.

